# Body fat volumes and distribution in chronic schizophrenia compared to healthy controls; a cross-sectional MR study

**DOI:** 10.1101/2021.09.20.21263820

**Authors:** Emanuele F. Osimo, Stefan P. Brugger, E. Louise Thomas, Oliver D. Howes

## Abstract

People with schizophrenia show higher risk for abdominal obesity than the general population, which could contribute to excess mortality. However, it is unclear whether this is driven by alterations in abdominal fat partitioning. Here, we test the hypothesis that individuals with schizophrenia show a higher proportion of visceral to total body fat measured using MRI. We recruited 38 patients with schizophrenia and 38 healthy controls matched on age, sex, ethnicity and body mass index. We found no significant difference in body fat distribution between groups, suggesting that increased abdominal obesity in schizophrenia is not associated with altered fat distribution.

## Introduction

Patients with schizophrenia show a 10-15 years shorter life expectancy (1), and their mortality due to natural causes is up to 8-times higher than expected (2). They have been shown to have an increased risk for abdominal obesity compared to the general population (odds ratio of 4.4 in a meta-analysis (3)). Abdominal obesity is the most prominent feature of the metabolic syndrome, and associates with blood lipid disorders, inflammation, insulin resistance or diabetes, and, downstream, increased risk of developing cardiovascular disease (4). Obesity, therefore, might underpin many of the metabolic comorbidities that are seen in schizophrenia, including a higher prevalence of hypertension, high cholesterol and triglycerides, type 2 diabetes, insulin resistance and the metabolic syndrome (CVD) (3). These, in turn, might be responsible for the increased cardiovascular mortality in this group (5).

Few studies have investigated adipose tissue mass and distribution in schizophrenia. Some have used proxies, such as bioelectrical impedance (BIA) (6, 7), while others have used gold standard MR techniques (8-10) to visualise body fat distributions in cases and controls with conflicting results. MR studies have reported no significant difference in visceral fat content between cases and controls (8-10), whereas studies using BIA have shown both increases (6) or decreases (7) in visceral fat in schizophrenia. However, studies measuring visceral fat using MRI tend not to measure total body fat content, and it is unclear whether these studies are able to detect differences in distribution at the whole body level in schizophrenia.

Our hypothesis was that patients and controls with schizophrenia, matched for body mass index, would show similar levels of total body fat, but an increased visceral fraction, as compared to controls.

## Methods

### Participants

Patients with schizophrenia were recruited from community mental health services in London, UK. Healthy controls (HCs) were recruited through the Hammersmith Hospital Healthy Volunteer Panel, London, UK, and through direct advertising, and were matched to patients for age (+/- 3 years), ethnicity, sex, and body mass index (BMI +/- 1).

Exclusion criteria for all participants were: age <18 or >65 years, pregnancy or breastfeeding, a history of cardiometabolic disease, including diabetes, hypertension, dyslipidaemia, ischaemic heart disease, any vascular disorder, other history of congenital/structural cardiac disease; or history of significant or continuing substance abuse. Inclusion criterion for patients was an ICD-10 diagnosis of schizophrenia. Exclusion criterion for healthy controls was a previous history or first-degree family history of schizophrenia or other psychotic disorder.

Written informed consent was obtained from all volunteers. The authors assert that all procedures contributing to this work comply with the ethical standards of the relevant national and institutional committees on human experimentation and with the Helsinki Declaration of 1975, as revised in 2008. All procedures involving human subjects were approved by the London - Camberwell St Giles Research Ethics Committee.

In addition to reviewing medical records, all subjects received a medical review and clinical examination to exclude medical co-morbidities. Body mass index (BMI) was calculated as body mass (kg) divided by the square of body height (m^2^).

### Magnetic Resonance Imaging

MR imaging was performed at a single site for all patient and controls. All participants but 29 HCs were scanned on a 3T Siemens Magnetom Prisma (Erlangen, Germany) using a combination of a 18-channel body coil and 12 elements of a 32-channel spine coil. An additional 29 whole-body fat imaging datasets were obtained at the same scanning site from HCs using a 1.5T Phillips Achiva scanner (Phillips, Best, the Netherlands). Previous QA measures comparing whole-body fat imaging datasets from individuals scanned on both 1.5T and 3T scanners demonstrate the data could be combined. A study of within-scanner variability (reproducibility) and across-scanner variability (including both 3T and 1.5T scanners) found that the within-scanner repeatability (coefficient of variation = 2.9%) explained much of the overall reproducibility (CoV = 4.4%) for visceral fat volumes (11).

Fat content and distribution were determined as previously described (12). Briefly, subjects were scanned using a rapid whole-body T1-weighted MRI protocol. Images were analysed using SliceOmatic (Tomovision, Montreal, Quebec, Canada) and regional volumes were recorded in litres (L), including total adipose tissue and visceral fat (13).

### Statistical analysis

Differences among patients and controls were tested using χ2 tests for categorical variables, Kruskal-Wallis tests by ranks for non-normally distributed values, and analysis of variance (anova) for normally distributed measures. Effect sizes were calculated using Cohen’s *d* measure.

In all tests, a p value <0.05 (two-tailed) was taken as significant. Statistical analyses and graph plotting were performed in R.

## Results

38 patients, as well as 38 matched HCs, underwent whole body fat MR measures. As expected, due to the nature of case control matching there were no significant group differences for age, sex, ethnicity and BMI. A sample description is in **Supplementary Table 1**. The patients were also included in a study of cardiac function (14)

**Table 1** describes MR-derived fat measures according to diagnostic status. There were no significant differences between groups in total fat (*d*=0.00, 95%CI: -0.46–0.46; p=1.00), visceral fat (*d*=0.06, 95%CI: -0.40–0.52; p=0.79), or visceral to total ratio (*d*=0.17, 95%CI: -0.29–0.62; p=0.47).

**Table 1:** MR-derived measurements in patients with schizophrenia and matched healthy controls.

| Characteristic | Sample size<br>N schizophrenia, N HC | Schizophrenia<br>Mean (SD) | Healthy Controls<br>Mean (SD) | Statistical results | Effect size (Cohen's *d*;<br>95% CI) |
| --- | --- | --- | --- | --- | --- |
| Total body fat (L) | 38, 38 | 30.27 (15.9) | 30.29 (15.2) | $F(1, 74) = 0.00, p=1.0$ | 0.00; -0.46–0.46 |
| Visceral fat (L) | | 3.83 (1.86) | 3.72 (1.92) | $F(1, 74) = 0.07, p=0.79$ | 0.06; -0.40–0.52 |
| Visceral fat ratio<br>Visceral/total body fat | | 0.13 (0.04) | 0.13 (0.05) | $F(1, 74) = 0.52, p=0.47$ | 0.17; -0.29–0.62 |

SD: standard deviation; CI: confidence interval; p= p value.

## Discussion

In this work we investigate the body fat distribution of people with chronic schizophrenia compared to healthy controls matched for age, sex, ethnicity, and body mass index.

We show that subjects with schizophrenia do not show any differences in overall adipose tissue content or regional distribution, when compared to matched healthy controls. This compares to the presence of concentric cardiac remodelling and cardiac fibrosis in the same patient cohort (14, 15). Our results are consistent with two prior studies which showed no difference in abdominal fat in schizophrenia relative to controls, and extend the previous literature on the topic by using whole-body MR imaging, to show that both body fat content and distribution were not different between cases vs controls. Strengths of our study include excluding any history of cardiometabolic disease, including diabetes, hypertension, dyslipidaemia, and ischaemic heart disease. Our results contrast with studies using body impedance measures, which have reported significant increases (6) or decreases (7) in visceral body fat in schizophrenia. Differences in patient selection and matching, sample size, and differences in measurement technique may account for the differences in results. Our main limitation is that all patients we included were chronic patients taking second-generation antipsychotics (SGAs), mostly clozapine and olanzapine. SGAs are known to cause increases in body weight (16), and are potentially one of the causes of the increased prevalence of obesity in schizophrenia. However, we note that previous studies looking at abdominal fat mass differences between drug-naïve patients with a first episode of psychosis and matched healthy controls (8, 9) found no difference even before antipsychotic drugs were started. It is also possible that our negative findings might be down to a statistical type II error, however our sample size was among the largest of similar studies, and our effect size measures were very close to 0 for all outcomes.

The lack of differences in visceral fat we observed in a closely matched control-case study add strength to similar findings from previous MRI studies. Differences in the prevalence of cardiometabolic disease in schizophrenia cannot be explained by specific alterations in body fat partitioning. Prospective studies will be useful to determine whether disease-related biological factors play a specific role in cardiometabolic disease in schizophrenia, or whether they simply reflect the same disease processes found in the wider population.

## Data Availability

The data that support the findings of this study are available on request from the corresponding author, ODH. The data are not publicly available due to ethics constraints and the potential for breaching participant privacy.

## Acknowledgments, contributions and funding

EFO and SB contributed to the design of the study, coordinated data collection, and recruited and scanned participants. EFO performed data analyses, and drafted the manuscript. ELT supervised data analyses and contributed to the manuscript. ODH conceived and designed the study, supervised data analyses, and contributed to the manuscript. All authors have approved the final manuscript. This work was funded by a Clinical PhD Fellowship to Dr Osimo jointly funded by the National Institute for Health Research (NIHR) Imperial Biomedical Research Centre (BRC) and the Medical Research Council (MRC) London Institute of Medical Sciences (LMS); by the BMA Foundation for Medical Research (Margaret Temple (2020) grant) to Dr Osimo and Prof Howes. This study was also funded by grants MC-A656-5QD30 from the Medical Research Council-UK, and the NIHR Biomedical Research Centre South London and Maudsley Foundation NHS Trust to Prof Howes. The authors would like to acknowledge the fantastic help provided by Mr Ben Statton (Superintendent Research Radiographer), Ms Alaine Berry (Senior Research Radiographer) and Ms Marina Quinlan (Research Radiographer), who carried out the scans and provided great support by anonymising, storing and retrieving them as needed.

## Conflict of interest disclosures

Dr Osimo, Dr Brugger and Professor Thomas report no conflicts of interest.

Professor Howes has received investigator-initiated research funding from and/or participated in advisory/speaker meetings organized by Angelini, Autifony, Heptares, Janssen, Lundbeck, Lyden-Delta, Otsuka, Servier, Sunovion, Rand, and Roche.

