## Supplementary Table 1 for "Body fat volumes and distribution in chronic schizophrenia compared to healthy controls; a cross-sectional MR study"

### Supplementary Table 1: Sample Characteristics

| Characteristic | Schizophrenia | Healthy Controls | Test statistic and p value |
|---|---|---|---|
| Sample size | 38 | 38 | |
| Male sex, N (%) | 29 (76%) | 29 (76%) | $\chi^2$ = 0; d.f.=1; p=1 |
| White ethnicity, N (%) | 17 (47%) | 19 (50%) | $\chi^2$ = 0.1; d.f.=1; p=0.78 |
| Age, years mean (SD) | 40.2 (10.0) | 39.7 (10.2) | t = 0.2, df = 74, p-value = 0.84 |
| BMI (kg/m2), mean (SD) | 28.45 (6.07) | 28.53 (5.44) | t = -0.06, df = 73, p-value = 0.95 |
| Chlorpromazine Equivalent Dose (mg/day), median (IQR) | 359 (274) | 0 (NA) | NA |
| Duration of treatment (years), median (IQR) | 12 (12.5) | NA | NA |

d.f.: degrees of freedom; p= p value; KW $\chi^2$: Kruskal Wallis chi squared; SD: standard deviation; IQR: inter-quartile range; N/A: not available; BMI: body mass index.
